# Nurse involvement in health information technology design for digital nursing practice: a scoping review

**DOI:** 10.1101/2025.09.15.25335735

**Authors:** Francis K. Kobekyaa, Dominique Boswell, Reilly Sinclair, Farinaz Havaei, Tracie Risling, Kristen R. Haase

## Abstract

There is growing demand for user involvement in health information technology (IT) design to ensure that emerging technologies meet the needs and expectations of end users. This scoping review explored the state of nurses’ involvement in health IT design, focusing on the: (1) methods, frequency, capacity, and levels of involvement in the design process; (2) types of health IT systems nurses are included in designing; and (3) outcomes of nurses’ inclusion in the design process. This review followed the Joanna Briggs Institute methodology for scoping reviews. A systematic search was conducted in seven databases: MEDLINE (Ovid), Cumulative Index to Nursing and Applied Health Literature (CINAHL) (EBCOhost), Embase (Ovid), Scopus, Web of Science, Compendex Engineering Village and Institute of Electrical and Electronics Engineers (IEEE) Xplore Digital Library database platforms. A total of 3,364 studies were screened at the title/abstract phase. After initial exclusions, 632 articles were screened for full texts, 495 were excluded, resulting in a final set of 137 eligible articles. Included studies most frequently involved nurses as users or testers of the technologies – usually at the end stages of the design process. Most studies interchangeably used the concepts of human-centred design and user-centred design to guide the design process. Interviews, surveys, observations and think-aloud techniques were the most frequently used design methods to elicit nurses’ perspectives about health IT systems. However, it was unclear which methods or approaches were most effective in engaging nurses in the design process. No standardized or validated nurse engagement frameworks were reported. These findings highlight the need to explore the nature of nurses’ involvement in design, their preferences in engagement and specific contributions and roles in order to develop an evidence-based approach to guide nurses’ participation in the design process.

**Author summary:** Nurses are rarely involved in health IT design, particularly in the early design stages. Failure to involve nurses in early stages can lead to IT systems lacking specific nursing contents, functionalities, and technical features, which may affect the use of the systems for nursing work. In this study, we found evidence of nurses being included at the end stages of the design process. Despite majority of the studies focusing on health IT design and development, nurses were mainly involved as users or testers of the technologies. In other words, nurses were mainly engaged in usability testing activities, which typically occur at the end of the design process rather than upstream design phases. We found that the concepts of human-centred design and user-centred design used interchangeably. At the end of the design processes, nurses perceived the IT systems as acceptable and usable, and either integrated or willing to integrate into clinical practice. Nurses perceived satisfaction with the design process resulting from effective technologies highlight the importance of engaging them in every stage of the design process. Our findings suggest that nurses have potential to leverage their clinical expertise and practical understanding of nursing workflows to guide health care technology design processes.

## Introduction

Health information technologies (IT) are increasingly utilized in healthcare settings across Canada and globally [1, 2]. Health IT includes computerised systems (e.g., clinical decision support systems (CDSS), electronic health records (EHRs)) which are designed to facilitate documentation, storage, and sharing of real-time patient data among care providers and patients [3]. These health IT tools have evolved from tools with simple functions such as capturing administrative data, to meticulously designed functionalities capable of gathering and integrating patient data into one repository [4-6]. The data in health IT systems is secure and accessible for healthcare providers [7], including nurses.

Nurses utilize these health IT systems for both direct and indirect nursing care, including documentation, communication, and clinical decision-making purposes [3, 8-10]. For example, documentation often involves using a closed-loop barcode medication administration system [11, 12], a speech recognition tool to capture nurses’ notes [13] or typical computer systems that have a screen to visually present notes and a mouse and keyboard for data input. The primary goal of these tools is to re-engineer nurses’ work processes, increase efficiency, and improve patient care quality [14]. As the design of these tools become more complex and diversified, they often fall short of meeting nurses’ needs due to lack of user-engagement [6, 15]. Focusing design efforts from nurses’ perspectives is critical for facilitating successful technology adoption in producing desirable outcomes [15-17]. Involving nurses to gain deeper insight into their unique design needs and preferences is required to create functional, user-friendly systems that support existing workflows [8, 18, 19].

Nurse involvement refers to the direct engagement and active participation of nurses in health IT design [20]. Nurses may be involved in various stages of the design process including developing empathy, defining and producing a health IT prototype after cycling through several rounds of the ideation and testing phases [21-23]. The engagement process is iterative, and could involve leveraging expertise from both nursing and human-computer interaction (HCI) disciplines [24]. HCI is a multidisciplinary field that focuses on designing user-friendly computerized systems and the study of peoples’ interaction with such systems [25]. This disciplinary mix of expertise accelerates creativity in ensuring health IT design is human-centered [24, 26]. Human-centered design involves empathy and inclusivity in the creation of interactive health IT systems. More specifically, it involves users (e.g., nurses) in the design process, ensuring that health IT systems are contextualized and tailored to their needs and capabilities [24, 27]. Nurses can have the opportunity to influence the design in a way that relate to their practice in patient care [28]. Because nurses need to routinely use tools that align with individual patient values and expectations according to the Canadian Nurses Association Code of Ethics [29]. Involving nurses could incorporate this view of patient-focused care into the design process [30]. More importantly, nurses could support the design of health IT systems that are user-validated and relevant to nursing practice, by leveraging their deep understanding of health IT systems and what is required to make them work more efficiently [30, 31].

However, little is known about the methods, frequency, capacity, or the levels at which nurses are involved in health IT design, including the types of health IT systems they design and the impacts of their involvement in the design process. Understanding these methods or levels of nurses’ involvement in health IT design is critical for meaningful engagement for efficient digital nursing practice and improved patient care. Therefore, this study sought to provide global evidence on the approaches, methods, frequency, capacity, and levels of nurses’ involvement in health IT design for nursing practice.

## Methods

Given that nurse involvement in health IT design process is an emerging area of research with little evidence synthesis, a scoping review was selected as the appropriate approach. This review was conducted following the Joanna Briggs Institute (JBI) methodology framework for scoping reviews [32]. The framework guided the researchers to: (1) identify and align the research question(s) and objective; (2) develop and align the inclusion and exclusion criteria with the review objective and question(s); (3) search for relevant evidence; (4) select the evidence based on the inclusion and exclusion criteria; (5) extract the evidence; (6) analyze the evidence; and (7) summarize and present the evidence in relation to the purpose of the review, making conclusions, and noting any implications related to the findings [32]. This scoping review was registered in the Open Science Framework database (Doi.org/10.17605/OSF.IO/53NH7).

### Review questions and objectives

The research questions that guided this scoping review were three-fold: (1) what is/are the approach(es), method(s), design focus, participatory activity(ies), frequency and/or levels at which nurses are involved in health IT design process? (2) what kind of health IT systems nurses are involved in designing? and (3) what are the outcomes of nurses’ involvement in health IT design? We addressed these questions through summarizing and analyzing global evidence on nurse involvement in health IT design for nursing practice.

### Inclusion and exclusion criteria

We used the broad: Population, Concept, and Context (PCC) framework recommended by JBI for Scoping Reviews [32] to determine the inclusion and exclusion criteria, as indicated in Table 1.

**Table 1:** Eligibility criteria based on Population, Concept, and Context framework.

| Determinants | Description |
| --- | --- |
| Population | Nurses of all specialties and categories, any age, any gender |
| Concept | Nurse involvement in health IT design for nursing practice. |
| Context | Healthcare settings such as inpatient, outpatient, community or virtual settings where people visit for healthcare services, worldwide. |

Studies were included if they: (1) Involved nurse(s) from any specialty, age or gender; (2) Related to eHealth or health IT design process (needs assessment/empathy, define, ideate, prototype, test/evaluate); (3) Conducted in healthcare settings (inpatient, outpatient, community or virtual) around the globe; (4) Any study design: quantitative, qualitative, and mixed methods design; and (5) Peer-reviewed studies published in English on/before April 9, 2025. Studies were excluded if: (1) nurses were not involved in the health IT design process; (2) technologies focused narrowly on physiologic monitors, intravenous (IV) pumps or teaching/learning purposes; (3) conference abstracts, review studies, theses or dissertations; and (4) not available in English or did not have their full text available, and published after April 9, 2025.

### Search strategy

The search strategy aimed to locate published articles. We conducted an initial pilot search on MEDLINE (Ovid) to identify relevant articles, assess whether a synthesis had already been completed, and uncover additional concepts to guide the final search strategy. The text words contained in the titles and abstracts of relevant articles, and the indexed terms used in describing the articles were noted and retained to assist in developing the full search strategy. This preliminary review helped us establish the inclusion-exclusion criteria, inductively create the first list of key words, and refine our research questions. Working with health sciences librarians (KM and EU), we developed a comprehensive final search strategy with keywords developed from MeSH databases and EMTREE for appropriate subject headings, text words and appropriate Boolean operators specific to each database. Given the multi-disciplinary nature of the study topic, we conducted a database search in nursing, medicine, and computer/engineering science-related databases including MEDLINE (Ovid), CINAHL (EBCOhost), Embase (Ovid), Scopus, Web of Science, Compendex Engineering Village, and IEEE Xplore Digital Library. Manual hand searching was conducted on April 9, 2025 to identify additional articles not retrieved through a database search.

### Evidence of selection and screening

Following the database search, all identified citations were collated and imported into Covidence systematic review software [33] on April 9, 2025. As a function of the Covidence software, duplicate articles were automatically removed. We further excluded articles that were reviews; posters; protocols; retracted papers; nurses/users not involved in design; educational health IT systems; no design/prototyping; health IT implementation, not designed for nursing practice, those with insufficient information; or that we were unable to access the full text. The authors of those articles that were not available in full text were contacted via email; however, none of the authors replied. We only retrieved one article through an interlibrary loan. The first three authors (FK, DB, and RS) screened all titles and abstracts, and full text reviews were conducted by FK and DB.

### Data extraction

Relevant data from the included articles were coded and extracted by three independent reviewers (FK, DB, and RS). Using an Excel spreadsheet data extraction tool developed by the authors, two reviewers (FK and DS) identified and categorized relevant themes based on the research questions in accordance with the JBI guidelines for data extraction [34]. The following data were extracted into the Excel sheet: author; year of publication; study setting; country of origin; title of the study; type of health IT; design focus; participatory design activity; frequency of nurse involvement; levels of nurse involvement; sample and category of nurses involved; nurses’ socio-demographics; design approach and methods; research approach and methods; theoretical frameworks/models; design guidelines, and key outcomes of nurse involvement in health IT design.

The three reviewers (FK, DB, and RS) conducted data extraction – each article was reviewed and extracted by two authors and then checked for consistency. The initial extracted data were discussed among all the three reviewers to ensure consistency, and then, for the approach was applied to the remaining papers.

### Data analysis

To answer the three research questions, the data extracted from the included studies was analyzed using descriptive qualitative content analysis [35]. Due to the heterogeneity of the data, meta-analysis was not possible. In accordance with the JBI methods of qualitative data synthesis and content analysis, codes were inductively generated and assigned to the highlighted texts or codes for each research question by two reviewers (FK and DB) [32]. Then, related codes were grouped into themes in line with the research questions [36]. Finally, the themes were reviewed by all authors and disagreements arising between reviewers were resolved through consensus discussion and/or with a third reviewer (KH).

## Results

Our comprehensive search yielded 13,652 articles and all were exported to Covidence systematic review software with 10,286 duplicates removed. From 3,366 articles, 2734 (93%) were excluded during the title and abstract screening phase. The remaining 632 articles were subjected to full text screening, resulting in a total of 137 articles tagged as ‘high relevance’ and therefore included for final analysis and synthesis as illustrated in the Preferred Reporting Items for Systematic Reviews and Meta-analyses extension for Scoping Reviews (PRISMA-ScR) [37] (see Fig 1).

**Fig 1.**
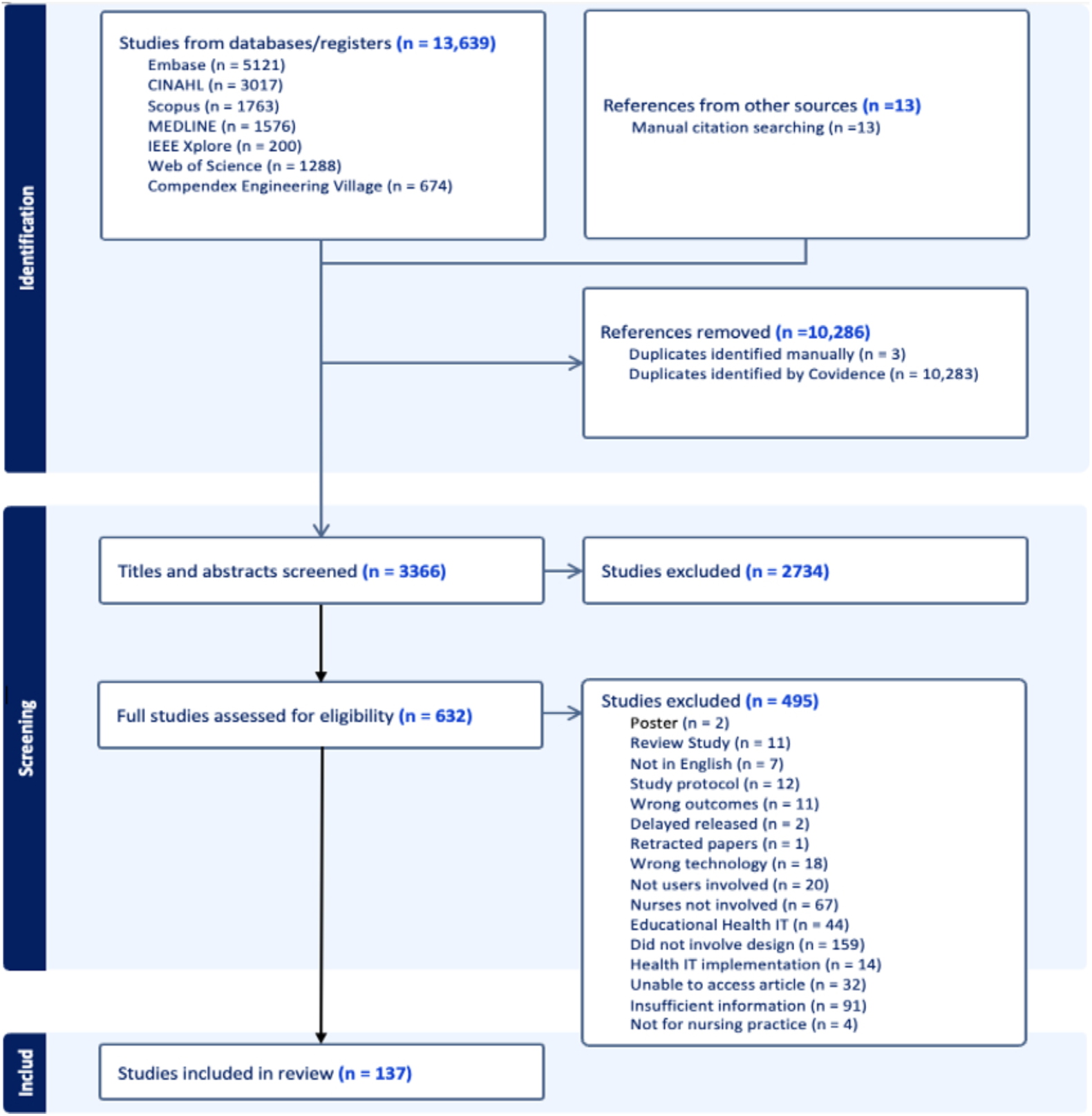
PRISMA-ScR flow diagram breaking down the selection of studies.

### Characteristics of included studies

The majority of the studies (130/137, 95%) were published between 2009 and 2025. Of all studies, 54 (39%) originated in North America with 48 from the United States and 6 from Canada [15, 38-87], 37 (27%) from Europe [88-123], 23 (17%) from East Asia [124-141], 9 (7%) from South America [142-150], 8 (6%) from Africa [69, 151-157], 4 (3%) from the Middle-East [158-161], and 2 (1%) from the Oceania or Australasia [162, 163] (Fig 2a).

**Fig 2a.**
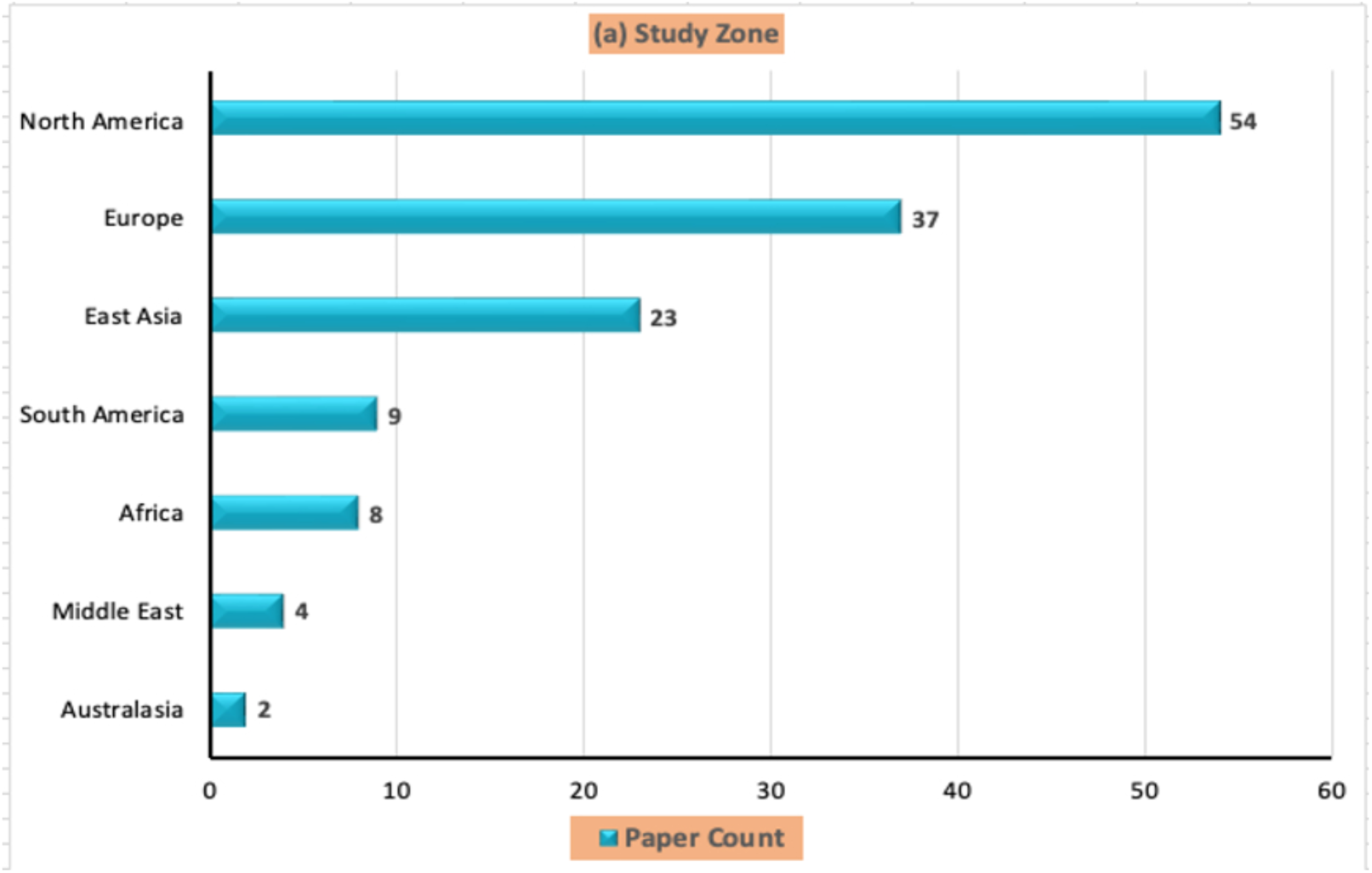
Characteristics of included studies, showing distribution of data by study zone identified for lead author(s).

Studies were carried out in different healthcare settings: 69 (50%) studies in general hospitals with undisclosed care locations [41, 42, 44–46, 51–56, 73, 76–82, 86–92, 95, 97–99, 107, 110, 111, 113, 114, 119, 122, 124–128, 130, 134–136, 139, 140, 142, 145, 147, 150, 153, 156, 157, 159, 161, 163-176], 25 (18%) studies in tertiary medical facilities [15, 38, 39, 59, 65, 67, 68, 71, 85, 94, 96, 101, 118, 120, 121, 123, 129, 141, 143, 144, 146, 148, 149, 154, 160], 11(11%) in home care settings [40, 57, 62, 64, 69, 73, 100, 130, 137, 162], 8 (8%) were in emergency departments [48, 61, 63, 93, 106, 108, 109, 112], 8 (8%) at cancer care centers [46, 47, 66, 70, 151, 152, 177] and 6 (6%) in intensive care units [72, 102, 115–117, 132]. Seven (5%) studies were conducted in simulation laboratory settings [49, 50, 60, 75, 83, 84, 104], and 3 (2%) in virtual settings [58, 105, 155] (Fig 2b).

**Fig 2b.**
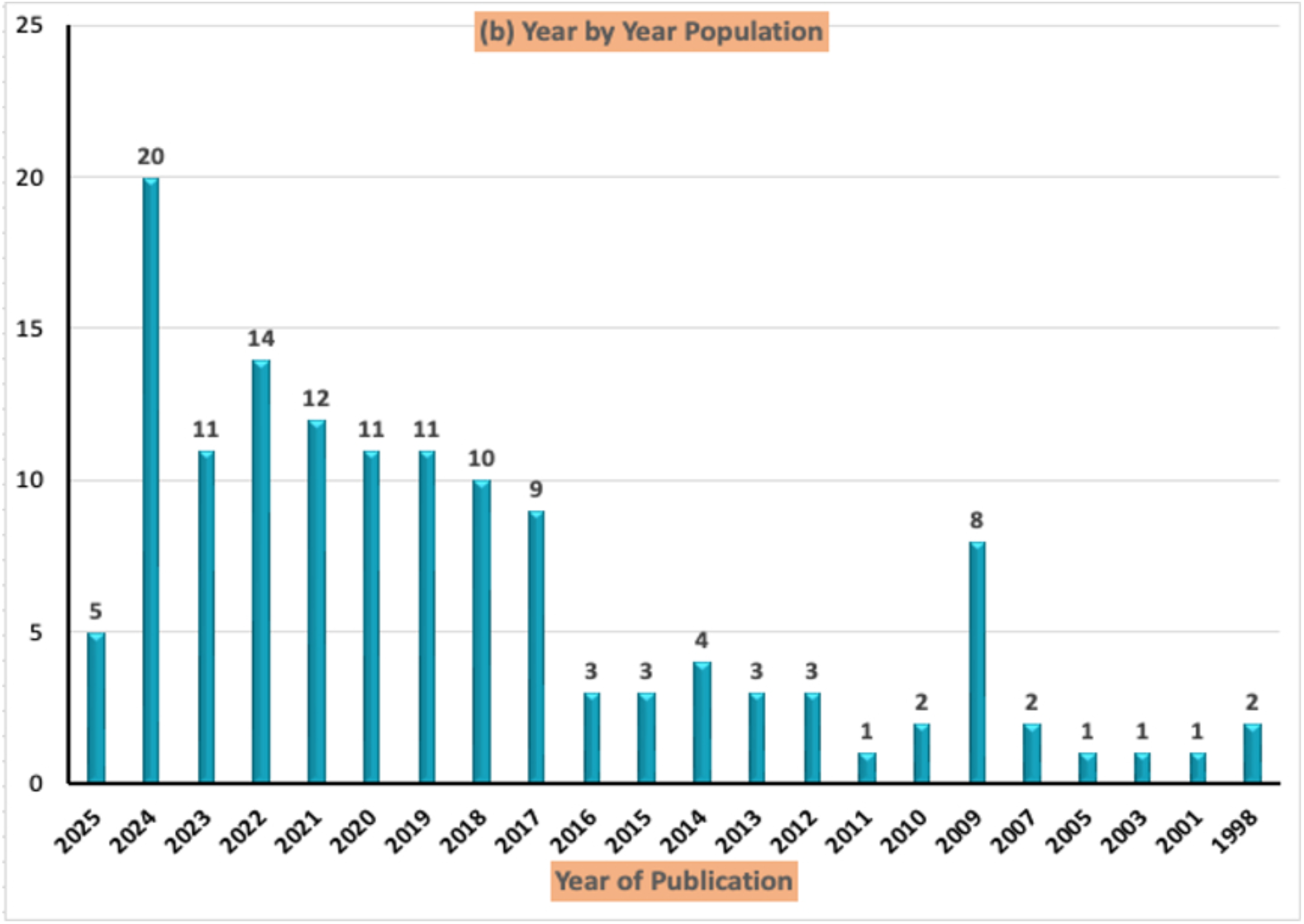
Characteristics of included studies, showing distribution of data, year by year publication – 1998 to 2025.

### Health IT design focus

The included studies addressed different components of health IT design processes. While some studies (13%, 18/137) focused on the entire design process, others addressed only some aspects of it. For example, 47 (34%) studies focused on usability testing of prototypes [60, 61, 67, 69, 70, 72, 73, 75, 76, 78, 83, 84, 86, 100, 104, 107, 108, 110, 112, 113, 116, 123, 143, 150, 169, 173, 175, 176] or evaluation and feasibility study of fully functional health IT tools [44, 50, 55, 57, 61, 107, 108, 121, 135, 140, 154, 164, 166, 169, 174], whereas others targeted evaluating health IT acceptance and adoption [60, 102, 110, 128, 157]. Additionally, 27 (19%) studies focused on designing and developing health IT tools or their prototypes/mock-ups [38-40, 42, 43, 68, 88, 91, 92, 99, 106, 111, 117, 126, 130, 137, 142, 146, 163, 178]. For example, Anderson et al. [38] designed a prototype for stroke prevention, while Chan et al. [39] focused on designing a prototype of a decision aid that could support decision making for hypospadias surgery. Other studies also focused on redesigning already existing health IT tools aimed at modifying or improving the software user interfaces [147, 153].

Twenty-six (18%) studies targeted primarily at eliciting technical requirements [15, 54, 58, 59, 62, 64, 81, 82, 87, 90, 103, 105, 124, 152, 159] or user requirements (i.e., needs and preferences) to guide design or programming of health IT applications [47, 49, 63–65, 77, 89, 98, 120, 149, 172]. For example, Chang et al. [124] gathered user needs for the design of a clinical decision support system for wound care management, whereas Busse et al. [89] conducted needs assessment for the design of an electronic cross-facility health records prototype.

Furthermore, 19 studies (13%) focused on health IT design, development, deployment, and user testing [45, 48, 62, 85, 94, 95, 118, 119, 125, 127, 129, 134, 136, 148, 151, 155, 156, 158, 161, 171] or needs assessment and usability testing only [97, 115]. More importantly, 18 (13%) studies addressed all aspects of health IT design ranging from needs assessment to user evaluation of health IT systems [46, 51–53, 56, 79, 80, 93, 96, 101, 116, 122, 131, 132, 141, 144, 145, 160, 163, 170]. For example, Ehrler et al. [93] identified functional specifications for an app programming and subsequently designed an app prototype, which was iteratively tested and evaluated through user interviews. Also, Novak et al.[179] engaged nurses to identify their needs and design requirements for a system design. The nurses and other members of the design team co-created a conceptual model based on the design requirements to guide the design of a low-fidelity prototype which was subsequently iteratively tested and refined. By involving nurses in the design process, the prototype developed was perceived to reflect their needs.

### Type of health IT system

Different types of health IT tools with diverse functions were identified across the included articles. The majority were unspecified eHealth tools or EHR systems (24%, 34/137) [43, 47, 50, 58, 67, 68, 75, 76, 78, 81, 88, 89, 106–108, 111, 114, 116, 126, 128, 134, 138, 144, 152, 157, 158, 160, 172, 175, 180-184] and clinical decision support systems (26%, 36/137) [38, 39, 45, 48, 49, 52, 56, 60, 62, 65, 70, 71, 82–84, 87, 95, 100, 102, 118, 120, 124, 127, 140–142, 145, 146, 154, 161, 163, 169, 173, 176, 185, 186]. Others were mHealth software or mobile health (mHealth) applications (16%, 23/137) [77, 80, 86, 92, 93, 98, 112, 119, 121, 122, 125, 136, 137, 148–151, 153, 164, 170, 171, 174, 187], nursing information/communication systems (10%, 14/137) [15, 41, 46, 64, 85, 94, 96, 99, 109, 135, 147, 162, 188, 189], electronic medication administration records (6%, 9/137) [44, 54, 55, 168, 190-194], web-based or online (virtual) applications (5%, 8/137) [51, 89, 91, 103, 104, 130, 155, 195], clinical dashboards (7%, 10/137) [40, 47, 53, 57, 63, 79, 97, 123, 156, 166] and remote monitoring systems (2%, 3/137) [42, 59, 105] (see Fig 3).

**Fig 3.**
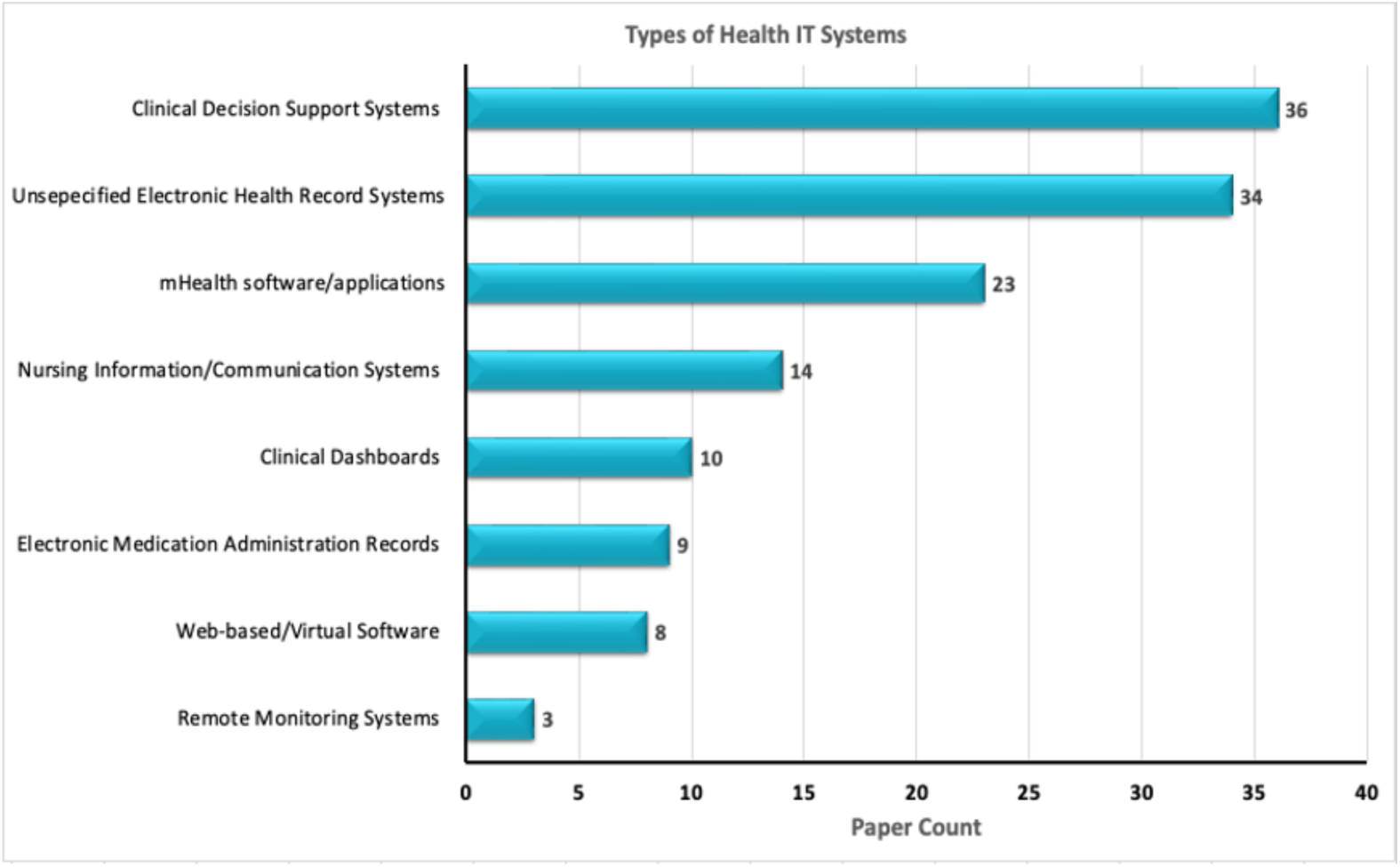
Type of health IT being designed and developed with nurses across the studies.

### Characteristics of nurses and their categories involved in the studies

Of all the included studies, 47 (34%) involved only nurses [15, 39–42, 46, 48, 54, 55, 58, 60, 62, 63, 67, 80, 82, 83, 85, 94, 98, 99, 102–105, 109, 111, 124, 128, 133–135, 140–142, 147, 148, 154, 160, 162, 164, 173, 183, 184, 188, 189, 191], with two health IT projects specifically designed and executed *by* nurses, *for* nurses [126, 127]. The majority (94%, 129/137) of the nurse participants in the projects were direct care nurses, with only 5% (8/137) done with nurse leaders [49, 62, 64, 100, 111, 187, 190, 195].

We also found studies involved nurses from several nursing specialities in the design process: 5 studies were done with nurses from intensive care unit [55, 63, 141, 188, 190]; 4 studies involved emergency nurses [48, 93, 112, 181]; 3 studies conducted with oncology nurses [46, 47, 121]; 3 studies with nurse anaesthetists [53, 56, 79]; and 2 studies with nurse informatics specialists [71, 127]. Other studies were conducted with nurses from other contexts: psychiatric/mental health [134]; cardiology [95]; surgery [119]; public health nurses [64]; pediatrics [125]; and home care units [105]. Of these care settings, some studies involved nurses from more than one setting in a single study [43, 46, 47, 49, 62, 181, 190, 192]; however, the majority (76%, 105/137) did not report on the specific care setting [15, 38, 39, 41–45, 50–52, 54, 57–60, 62, 65, 67–70, 75–78, 80–90, 92, 94, 96–100, 102–104, 106–109, 111, 114, 116, 118, 120, 122–124, 126, 128, 130, 133, 135, 136, 138, 140, 142, 144–158, 160–164, 168–176, 182–184, 186, 189, 191, 193-195]. Seven studies reported involving nursing professionals with technical and higher education levels, computer experience, including their age and gender [55, 61, 66, 70, 89, 103, 112, 128, 130, 135, 143, 151, 196-200], as it was perceived that nurses’ personal characteristics could influence their levels of engagement in health IT design. It was also perceived that nurses past experience with technology use is linked to their readiness to accept them after deployment [128].

Most studies involved between 1-20 nurses (see Table 2); 14 (10%) of the studies did not report on the number of nurses that participated in the various design stages [39, 41, 59, 81, 104, 105, 111, 126, 142, 150, 156, 163, 170, 172].

**Table 2.**
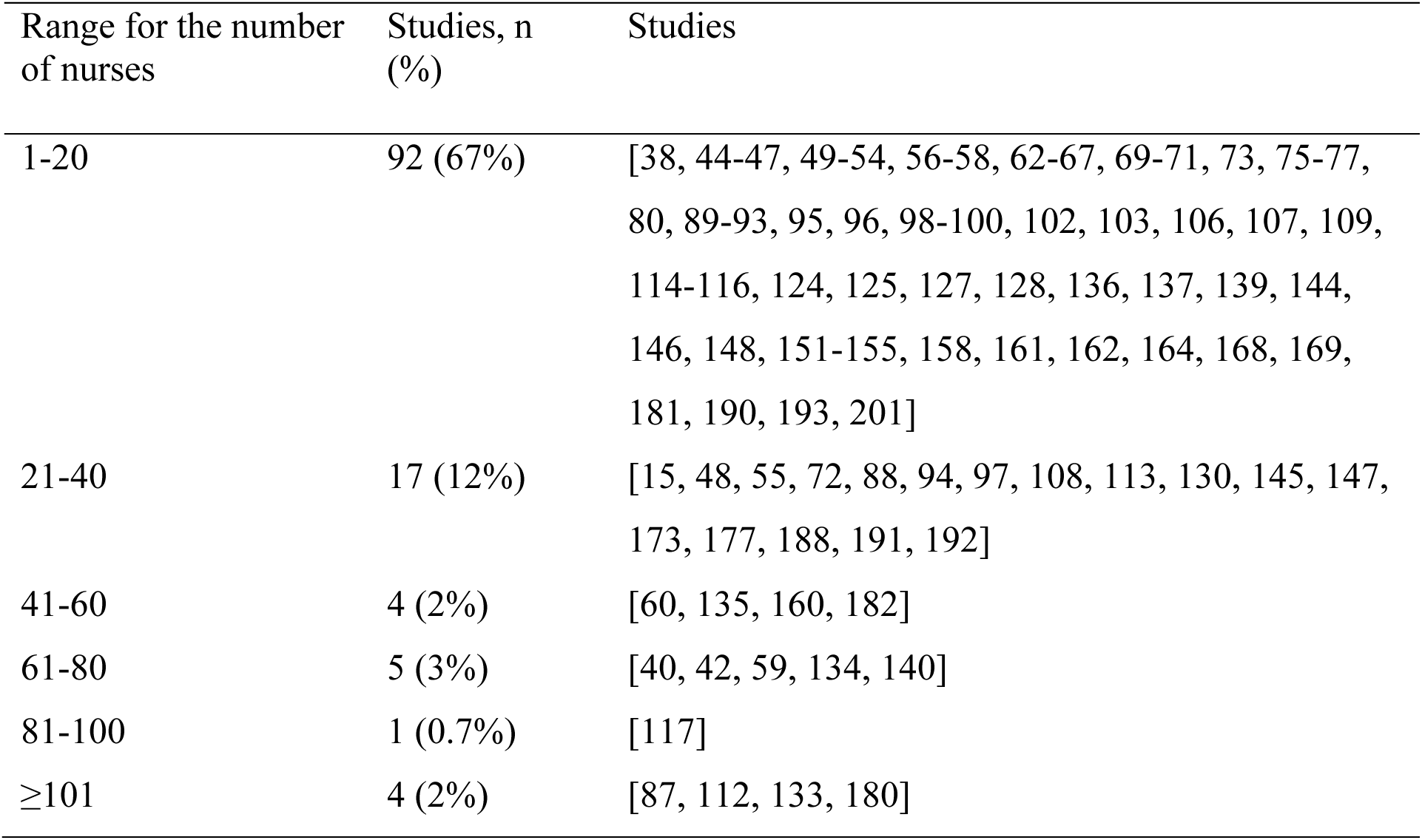
Average number of nurses per study (n=18)

| Range for the number of nurses | Studies, n (%) | Studies |
| --- | --- | --- |
| 1-20 | 92 (67%) | [38, 44-47, 49-54, 56-58, 62-67, 69-71, 73, 75-77, 80, 89-93, 95, 96, 98-100, 102, 103, 106, 107, 109, 114-116, 124, 125, 127, 128, 136, 137, 139, 144, 146, 148, 151-155, 158, 161, 162, 164, 168, 169, 181, 190, 193, 201] |
| 21-40 | 17 (12%) | [15, 48, 55, 72, 88, 94, 97, 108, 113, 130, 145, 147, 173, 177, 188, 191, 192] |
| 41-60 | 4 (2%) | [60, 135, 160, 182] |
| 61-80 | 5 (3%) | [40, 42, 59, 134, 140] |
| 81-100 | 1 (0.7%) | [117] |
| ≥101 | 4 (2%) | [87, 112, 133, 180] |

### Levels of nurse involvement

To understand the levels of nurse involvement, we coded the studies based on Druin’s framework [202], which involves four levels and roles of technology design as: users, testers, informants, and design partners (see Fig 4). The majority (33%, 46/137) of the studies involved nurses as testers in the design process [40, 42–45, 50–52, 60, 61, 63, 66, 67, 70, 72, 73, 76, 86, 89, 92, 95–97, 99, 106–108, 112, 113, 129, 130, 135, 136, 141, 144, 146, 150, 154, 173, 189, 200, 203-205]. As testers, they evaluated and offered feedback on prototypes of health IT software before final development and deployment for use in clinical practice. The results of the testing were meant to redesign [153, 206] or modify and/or improve future iterations of the prototypes before they were implemented [44, 47, 107, 113, 154]. We also found 32% (44/137) of the studies engaging nurses as informants [15, 39, 41, 47, 53, 55, 58, 62, 64, 65, 68, 69, 71, 79–82, 87, 91, 103, 105, 109, 114, 116, 122, 125, 127, 128, 132, 142, 143, 148, 149, 151, 153, 160, 165, 172, 206-212]. In the role of informants, nurses were engaged in the design process at different phases, sharing ideas during brainstorming, and providing input and feedback to design prototypes.

**Fig 4.**
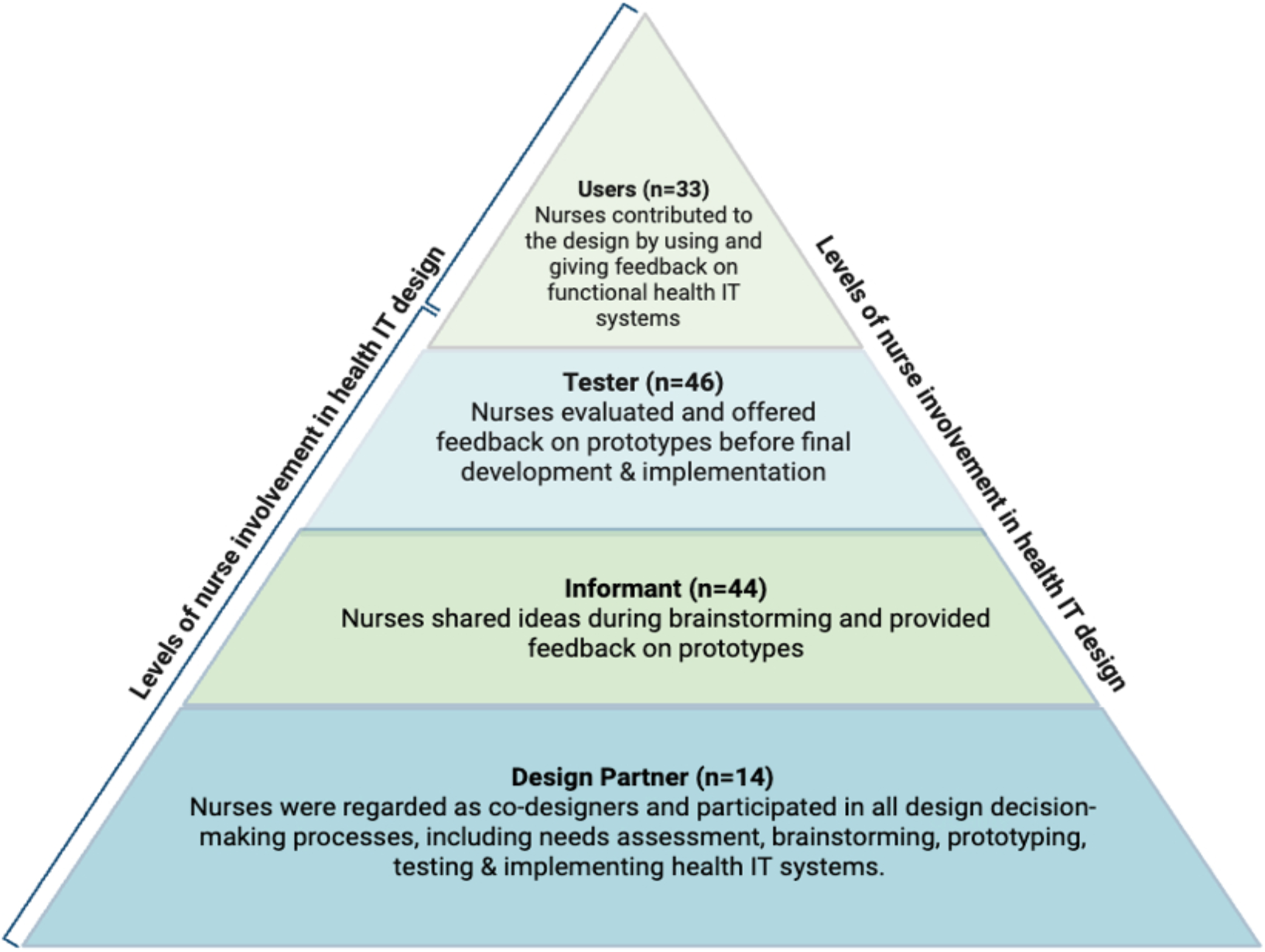
Levels of nurse involvement in health IT design process according to Druin’s framework.

Moreover, as users, 33 (24%) of the studies engaged nurses to contribute to the design by using and giving feedback on functional health IT tools [47, 54, 56, 77, 78, 83, 84, 90, 98, 101, 104, 120, 121, 123, 124, 133, 140, 155–157, 159, 161, 163, 168, 170, 171, 174–176, 197–199, 213, 214]. Lastly, 14 (10%) studies included nurses as design partners, where they were regarded as equal partners or co-designers in the design process, directing and influencing decision-making processes in all phases of the design process – from needs assessment to testing and evaluation [38, 49, 57, 85, 94, 100, 115, 117–119, 126, 138, 158, 215].

### Frequency of nurse involvement

The frequency of nurse involvement was coded based on the number of times they were engaged in the design process (Fig 5a). While some studies involved nurses at several points across the design process or had repeated engagements, others engaged them only one time. For example, in 40% (55/137) of the studies, nurses were engaged once [40, 41, 44, 54, 59, 60, 63, 64, 66, 69, 73, 76, 77, 82, 90, 98, 102, 104, 105, 107, 109, 110, 112, 113, 119, 122, 123, 128–130, 132, 133, 135, 136, 138–141, 143, 146, 149–154, 157, 159, 161–164, 168, 169, 173]. In 30% (42/137) of the studies involved nurses two times [15, 42, 45, 47, 48, 51, 56, 57, 61, 62, 67, 70, 75, 80, 81, 85, 88, 89, 91–93, 97, 99–101, 106, 108, 114, 118, 120, 121, 127, 134, 137, 141, 144, 170, 171, 175, 176], 15% of the studies (21/137) involved nurses 3 times [46, 49, 50, 52, 55, 58, 79, 83, 84, 86, 95, 96, 125, 145, 148, 155, 156, 158, 160, 165, 172], 7% (10/137) reported 4 times of engagements [38, 53, 68, 71, 72, 103, 115–117, 147], and 5% (7/137) reported to have engaged nurses more than 4 times [43, 65, 71, 94, 111, 124, 142].

**Fig 5a.**
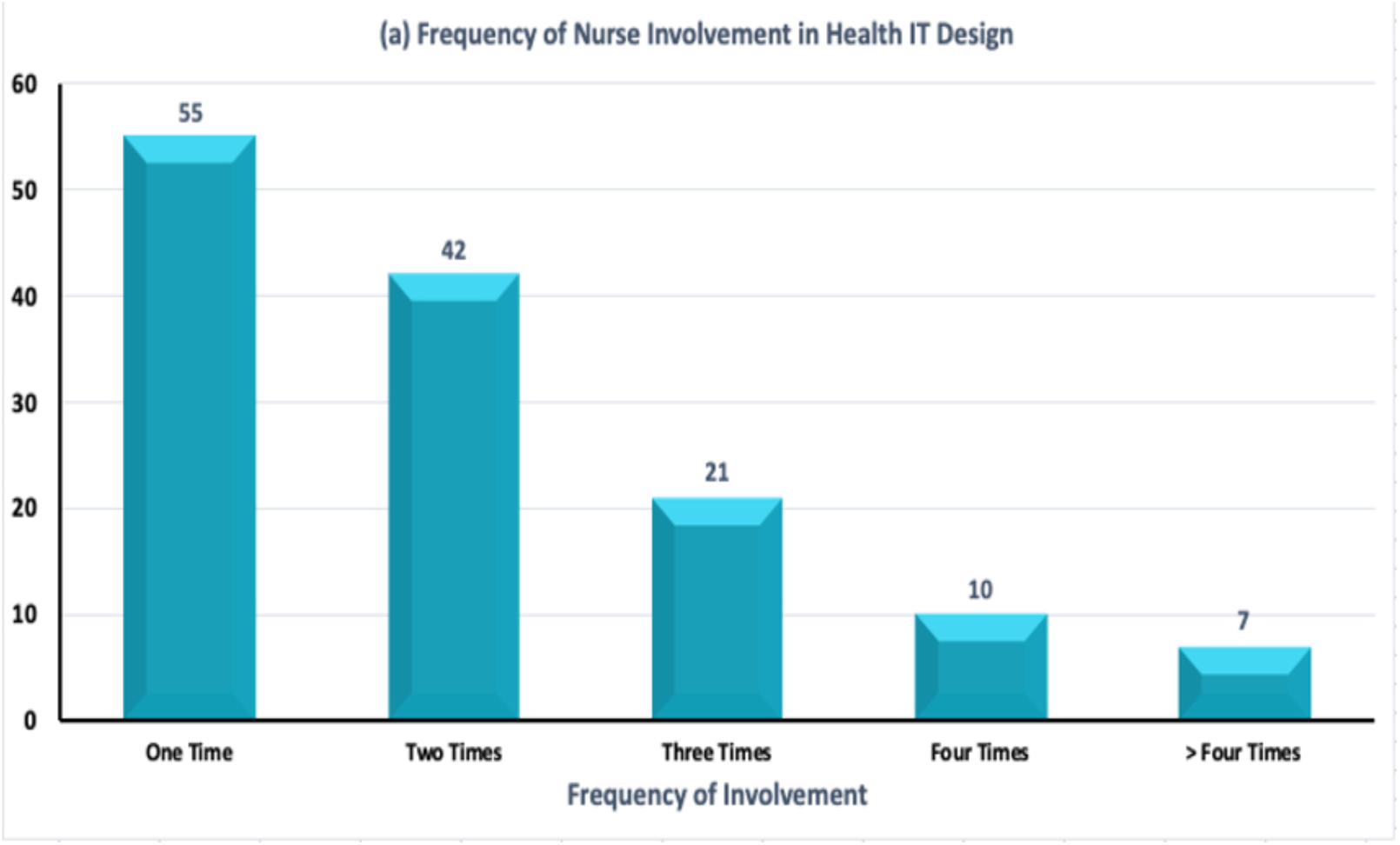
Nurse involvement in design, showing distribution of data by frequency of nurse involvement in the design process.

### Participatory design activities

The participatory activities nurses engaged in during the design process were analyzed (Fig 5b). In 53% (73/137) of the studies, nurses were involved in usability testing of health IT tools – usually at the later stages of the design process (i.e., formative/summative evaluation) [15, 40, 42-45, 47, 50, 51, 57, 60-62, 66-70, 72, 73, 76, 79, 84, 86, 90, 92, 104, 106-108, 112, 113, 118, 120, 121, 123, 125, 128-130, 132, 133, 135, 136, 140, 143, 145, 146, 150, 151, 153, 154, 156, 157, 161, 163, 170, 171, 173, 174, 176, 189, 198, 200, 203-206, 210, 211, 213]. This was followed by user or technical requirements gathering with 24% (33/137) [41, 47, 54, 58, 63, 64, 77, 78, 81, 82, 87, 89, 91, 95, 96, 98, 103, 105, 109, 119, 124, 127, 149, 158-160, 168, 197, 199, 207, 209, 214, 215] of studies reporting this and only 6% (9/137) noted involvement in the actual design through design workshops or prototype sessions [39, 71, 80, 83, 85, 101, 122, 126, 175]. In contrast, 14% (20/137) of the studies involved nurses across several components of the software development phases including needs assessment (i.e., defining user and design requirements, contents and functionalities) and interface evaluation [52, 53, 56, 97, 99, 114, 141, 144, 148, 165, 172, 208, 216] or all the phases combined [38, 49, 65, 94, 100, 115, 116]. In one study, nurses proposed the design idea and actively participated in every stage of the design process, including needs assessment, development, implementation, and evaluation of the health IT tool [117]. Similarly, in Guo et al.’s [126] study, nursing staff designed and developed for themselves an evaluation system using Microsoft in-house Excel Visual Basic for Applications (VBA) tools for managing dry skin in elderly patients in China. In 2 out of the 137 studies, we found that nurses were observed while using health IT tools for nursing care and interviewed afterwards to elicit their experiences [54, 55].

**Fig 5b.**
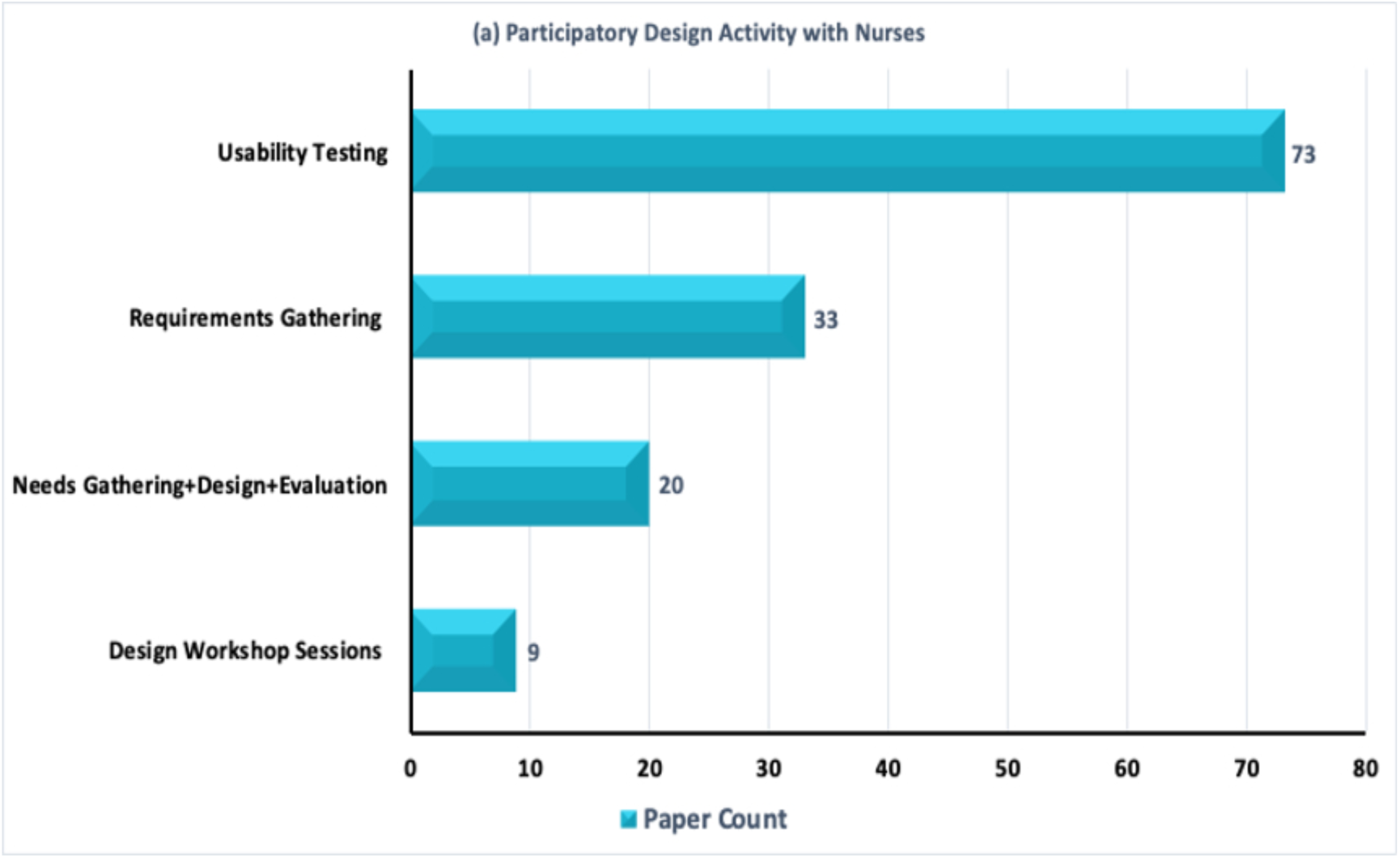
Nurse involvement in design, showing distribution of data by participatory design activity.

### Study methodologies, designs, and methods

Various research methodologies and methods as well as design techniques and methods were adopted to guide nurses’ involvement in health IT design. The majority of the studies (45%, 63/137) did not report on the research methodology [15, 40-42, 56-59, 61, 63, 67, 69, 71, 72, 75, 78, 79, 82-85, 94, 96, 97, 102, 103, 106, 108, 109, 111, 113, 115, 116, 118-120, 122, 126, 129, 130, 132, 136, 138, 142, 144, 146, 147, 149, 150, 153, 155, 158, 161, 163, 165, 168-171, 174, 189, 208, 215]. However, 27% (37/137) reported a qualitative approach to understand user needs or conduct usability testing [39, 41, 43, 47, 49, 51, 53, 54, 62, 64-66, 68, 70, 73, 87, 89-92, 98, 99, 104, 107, 114, 123, 124, 127, 154, 157, 159, 162, 197, 200, 207, 210]. Approximately 18% (25/137) adopted a mixed methods approach [38, 52, 55, 76, 77, 80, 81, 86, 95, 100, 101, 105, 117, 121, 141, 143, 145, 148, 152, 156, 164, 172, 175, 176], while 8% (12/137) adopted quantitative approach but were not explicit in their description [44, 45, 50, 60, 112, 128, 133-135, 151]. Quantitative approaches were often used to evaluate the usability or acceptability of health IT tools, or to support the qualitative findings [44, 60, 112, 140, 173].

Many of the studies also reported on a number of software design strategies. While 26% (36/137) of the studies did not report on any design methods [38, 44, 47, 52, 54, 60-63, 67, 73, 80, 90, 92, 101, 105, 108, 116, 118, 126-129, 133, 135, 140, 143, 158, 159, 168, 199, 203, 206, 210, 213, 216], 24% (34/103) utilized a user-centered design [46, 50, 53, 55, 56, 59, 68, 72, 77-79, 81, 83, 86, 87, 97, 99, 100, 112, 119, 124, 131, 144, 145, 148, 155, 156, 162, 174, 176, 185, 189, 198, 206], 13% (19/137) a participatory design [47, 49, 51, 58, 76, 95, 96, 103, 107, 109, 111, 114, 115, 132, 160, 161, 178, 197, 200], 8% (11/137) an iterative design [42, 43, 45, 57, 70, 71, 94, 113, 146, 151, 187] and 7% (10/137) a human-centered design [39, 82, 84, 91, 98, 104, 106, 157, 167, 208]. Other studies applied the software development life cycle [40, 136, 142, 173, 197], design science information systems and software/design science research methodology [150, 170, 175], work-centred design [74, 211], analysis, design, development, implementation, and evaluation (ADDIE) approach [121, 171], discipline agile delivery [120, 122, 149, 172], co-design [15], rapid application development methodology [123], multidisciplinary design [125], design thinking [153], double-diamond design framework [87] and Nielsen and Norman’s 4-phased design cycle [85]. Some studies employed more than one design methods [50, 64, 115, 130, 136, 154, 160, 163, 206, 207, 211]. These combined methods include user-centred design and software development life cycle [136], user-centered design with human factors engineering approach [141], participatory design, iterative and human-centred design as well as participatory and scenario-based design methods [64, 154].

We found that the concepts of human-centred design and user-centred design were used interchangeably and mostly referred to the application of the International Organization for Standardization (ISO) 9241-210 requirements [144, 148]. ISO 9241-210 standard is an international association of national standards bodies with the mandate to offer principles of human-centered design and ensure the application of the method in the life cycle of systems design [217]. However, human-centered design was among the least commonly found designs in the studies, though it somewhat enhanced nurses’ involvement in the design process [42].

A wide range of traditional research methods alongside methods specific to design were employed to engage nurses and other stakeholders in the design processes (see Table 3). The most common research methods were surveys (23%, 32/137) [38, 42, 45, 46, 50, 82, 95, 112, 113, 119, 122, 128-130, 133, 135, 140, 143, 144, 148-151, 158, 161, 171, 173, 198, 208, 213, 216], individual interviews (13%, 23/137) [47, 51, 56, 66, 83, 85, 87, 90, 100, 105, 108, 116, 123, 124, 127, 156, 157, 162, 167, 168, 203, 208] and focus group discussions (5%, 8/137) [39, 43, 53, 125, 132, 163, 199, 207]. 47% (65/137) of the studies combined surveys, interviews or focus group discussions with observations [15, 38, 40, 46, 52, 54, 55, 57, 60-64, 66, 67, 70, 72, 76, 77, 79-81, 84, 86, 89-92, 94, 96-101, 103, 104, 106, 108, 109, 118, 120, 121, 131, 145, 146, 153-155, 159, 160, 170, 172, 174-176, 185, 187, 197, 206, 207, 210, 211]. Employing these methods, the researchers observed nurses as they carried out nursing activities while using the health IT applications and subsequently conducted face-to-face interviews with them to authenticate their observations [40, 54, 61, 63, 72, 96, 97, 99, 106, 131, 153, 187, 206]. The observations were carried out in both clinical settings and simulation laboratories [218].

**Table 3.** Study methodologies, designs, and methods.

| Research Approaches | HCI Design Techniques | Research Methods | HCI Design Methods |
| --- | --- | --- | --- |
| Qualitative | User-centred design | Interviews | Think-aloud techniques |
| Quantitative | Human-centred design | Focus groups | Card sorting |
| Mixed Methods | Participatory design | Survey/Questionnaire | Task analysis |
|  | Iterative design | Observations | Personas |
|  | Multidisciplinary design | Delphi technique | Storytelling |
|  | Software dev't life cycle | Expert meetings | Storyboarding |
|  | Design thinking | Document reviews | User case scenarios |
|  | Design science info.<br>Systems and software |  | Heuristics evaluation |
|  | Discipline agile delivery |  | Cognitive walkthrough |
|  | Human-factors<br>engineering approach |  | Brainstorming/writing |
|  | Nielsen and Norman's<br>four-phased design cycle |  | Design workshops |

Out of the 137 studies, 63 used HCI design methods. Of these 63 studies, 21 (33%) applied concurrent think-aloud technique during usability testing [38, 53, 62, 66, 70, 73, 76, 79, 84, 91, 92, 97, 100, 125, 153, 156, 158, 176, 185, 198, 200]. In the think-aloud method, as participants interacted with the tool, they were asked to verbalize their thoughts and feelings about the software [70]. Nine (14%) studies conducted heuristic evaluations [40, 44, 57, 74, 100, 104, 115, 160, 198], 3 (6%) cognitive walkthroughs [62, 96, 108], and 7 (11%) employed user case scenarios or vignettes [38, 46, 49, 60, 62, 96, 148, 198]. Moreover, in 10 (15%) studies, design workshops were organised with nurses and other stakeholders to generate ideas related to design requirements, prototype solutions, and to provide feedback on designs [39, 83, 95, 103, 107, 111, 114, 145, 160, 219]. The least commonly used design methods were personas [62, 63, 153, 160], card sorting [15, 21, 86], storyboarding [81, 172], task analysis [40], brainstorming and/or brainwriting exercises [21, 160], storytelling [21], and document analysis [159]. We further found that 43 studies employed both research and design methods together [15, 38-40, 46, 49, 53, 57, 60, 62, 66, 68, 73, 74, 76, 79, 81, 83, 84, 86, 91, 92, 95-97, 100, 104, 108, 114, 115, 125, 148, 153, 154, 156, 158-160, 172, 176, 185, 198, 200].

### Theoretical frameworks and design guidelines

We explored theoretical underpinnings and design guidelines that provided foundations for the health IT design processes in the included studies (Table 4). Of the 137, Technology Acceptance Model (TAM) was used in 6 (4%) studies [46, 62, 128, 129, 157, 207], Unified Theory of Acceptance and Use of Technology (UTAUT) in 4 (2%) studies [89, 112, 197, 200], and Ottawa Decision Support Framework (ODSF) in 2 studies [39, 95]. Feedback Intervention Theory (FIT) in 1 study [40], Knowledge-to-Action Model and Duchscher’s Stages of Transition Theory [80], Phased Research Framework [87], and Information System Research (IRS) framework [119]. Other frameworks include: Framework for Societal Development Through Secure Internet of Things and Open Data (SSio) Health Care Services [174], Human-Centered Design Life Cycle Model [78], Family Nursing framework [118], Agile Development Model for Discovery, Development, and Deployment framework [172]. Several other frameworks were reported in the studies (see S4 Table); however, the majority (64%, 88/137) of these studies did not explicitly report or specify the frameworks used [15, 41-45, 47, 50, 53, 55, 56, 58, 60, 61, 63-67, 69, 70, 73, 76, 77, 79, 82-86, 90, 91, 96, 98-101, 103-109, 122, 123, 126, 133, 135, 138-141, 143-146, 149, 151, 154-156, 158, 159, 161, 163, 168, 170, 171, 173, 175, 176, 198, 203, 204, 209, 211].

**Table 4.**
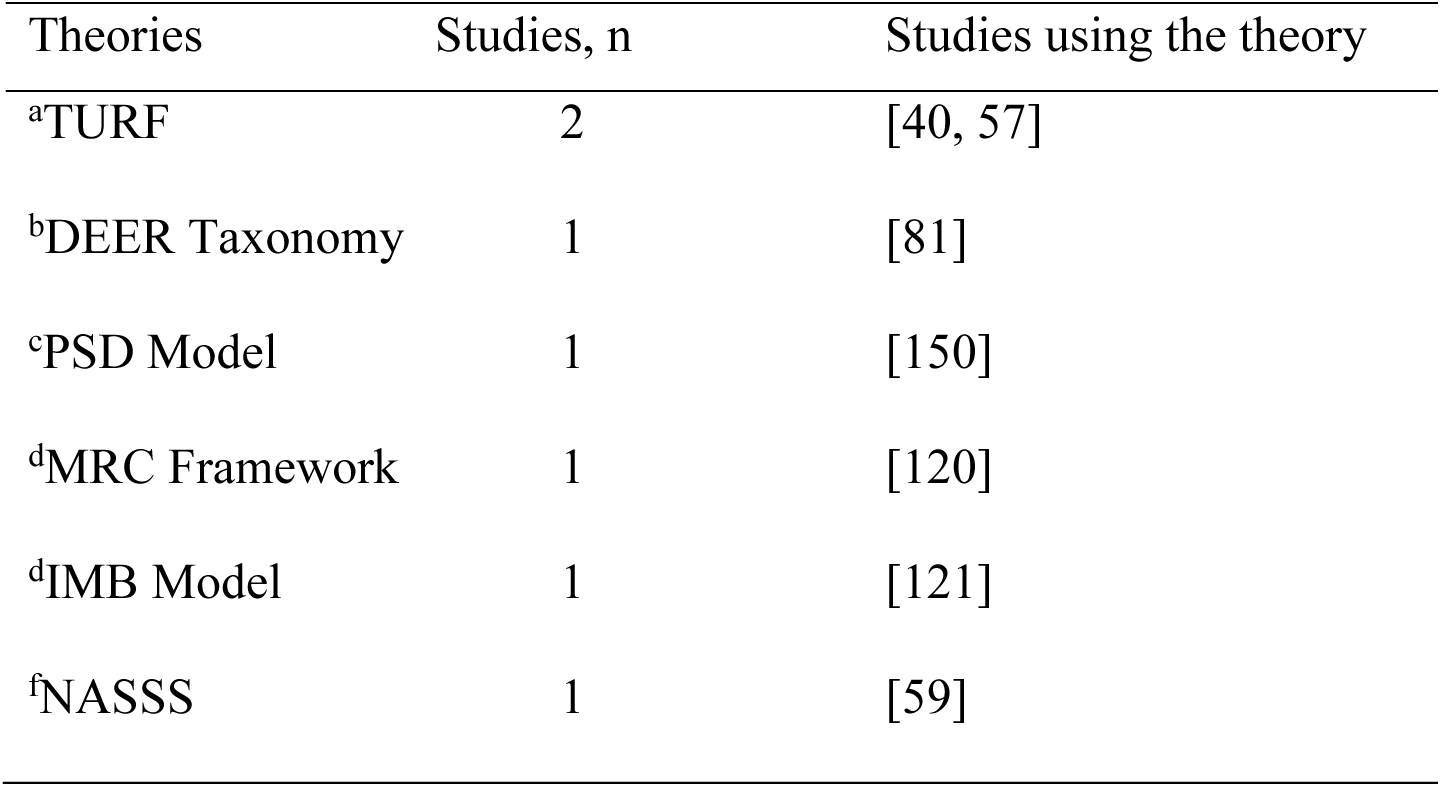
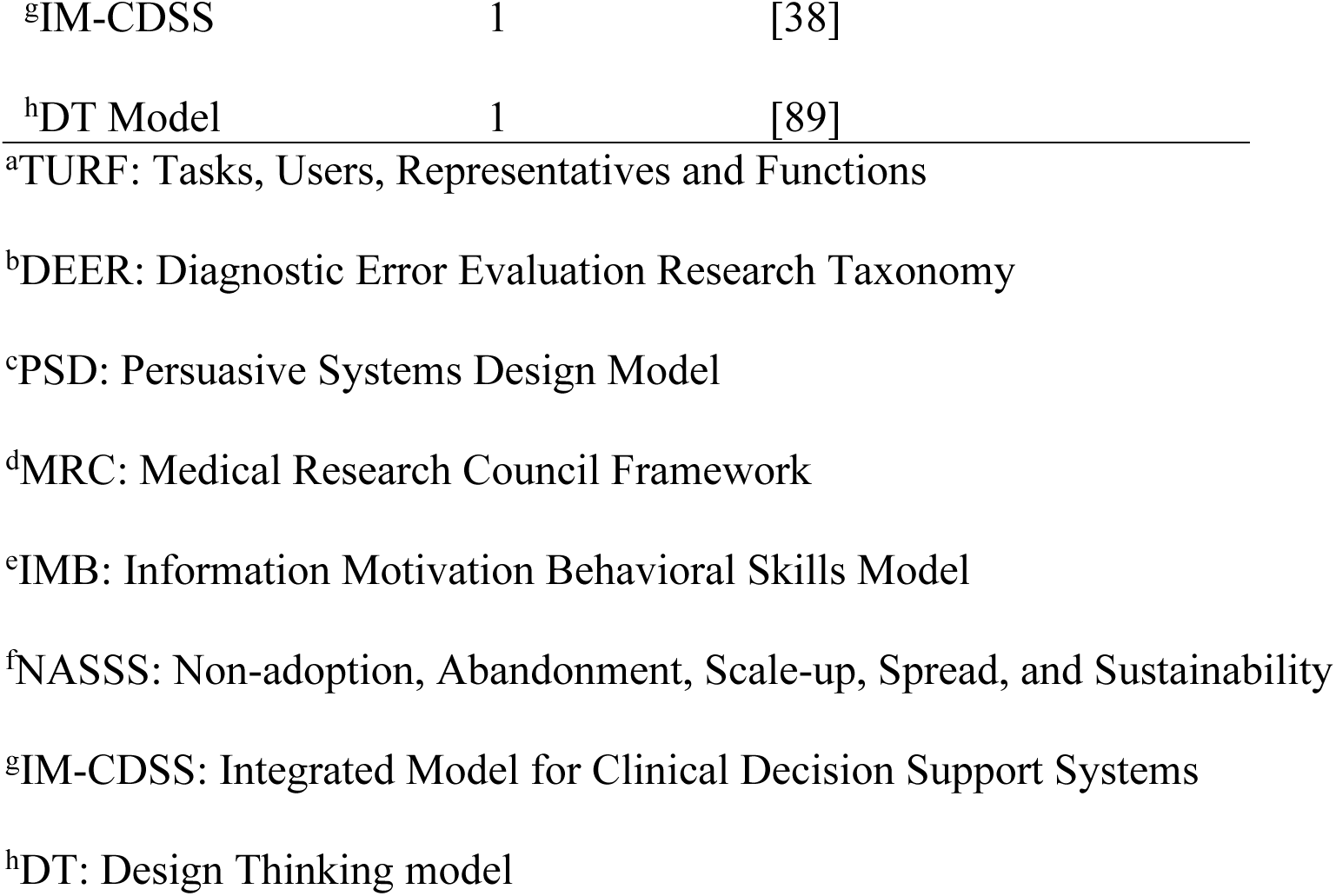
Major theories and design guidelines used, categorised with citations.

Furthermore, a total of 19 (13%) studies used design principles or guidelines to inform the design and evaluation processes. Of these 19 studies, 11 studies [40, 44, 57, 66, 100, 104, 108, 127, 143, 144, 148, 208] applied the 10 Nielsen’s heuristic principles Nielsen (1994a); 2 studies used the American Medical Informatics Association Usability design principles [140] and Human-factor Engineering (HFE) principles [141] respectively, to evaluate and diagnose design problems, optimize user interfaces, and to enhance user experience; six studies applied unspecified design principles to design user interfaces [57, 62, 103, 144, 146, 200]. The majority (84%, 87/103) of the studies did not report on any design principles applied in the designs.

### Outcome of the design

Although no studies utilized a validated instrument to assess nurses’ involvement in design, we coded the outcome of the design process based on the feedback reported by nurses. From a design perspective, the design of a new software, system or a prototype is considered an outcome [26]. Three categories were created to describe the outcome: (1) acceptability, adoption, and integration; (2) usability and utility; and (3) satisfaction. 32 studies reported that nurses perceived the health IT systems or their prototypes acceptable and either integrated or willing to integrate into clinical practice [38, 45, 52, 56, 60, 67, 81, 83, 91, 95, 96, 99, 104, 115, 116, 118, 120, 127, 129, 132, 133, 139, 140, 149, 157, 160, 162, 174, 206, 208, 210, 216]. Similarly, in 32 studies, nurses’ feedback enhanced the systems design and described the technologies as effective, useful and usable for nursing practice [15, 46, 53, 57, 61, 76, 78-80, 84, 86, 92, 105, 113, 119, 122, 125, 126, 128, 130, 141, 148, 150, 151, 155, 161, 170-172, 175, 207, 211], and 15% (21/137) found the technologies to be satisfactory, reflecting their nursing needs and improving patient care [77, 82, 85, 90, 97-99, 121, 123, 131, 135, 144, 145, 147, 156, 173, 176, 203, 220-222].

## Discussion

This scoping review provides global evidence regarding the state of nurse involvement in health IT design, focusing on the approaches, methods, design focus, frequency, and the levels or capacity at which they are involved. Nurse involvement in designing health IT systems is important given the potential of ensuring IT systems align with daily nursing routines [30, 223], improve workflow efficiency and optimize patient care [224]. Novak et al. [179] noted that involving nurses throughout the design process could help meet their needs. Because nurses have profound expertise and perspectives on what health IT platforms they need, and these perspectives can be taken and incorporated into health IT systems design with relative ease [26]. However, unlike prior research that emphasizes end-users inclusion in all phases of the design process [30], this research found otherwise. Rather than involving nurses early and in all phases of the design process to create and customize health IT systems that resonate with patient care [30], we found that nurses were mostly involved at late stages of the design process. At the late stages, nurses only participated in usability testing, evaluating how usable the systems were rather than engaging in the actual design sessions. For instance, in Ehrler et al.’s [207] study in Switzerland, nurses only participated in prototype evaluations but were excluded from the actual prototype design stage. This limited participation reflects tokenism reinforcing the marginalization of nurses’ voice in health IT design processes [30]. Our findings thus suggest that design teams need to recognize the importance of nurse involvement at project conception and throughout the design stages reflecting the need for inclusive design approaches [225]. Although inclusive design largely focuses on design with older adults and people with disabilities [225, 226], this focus could be extended to nurses – who are historically rarely being included in the design process [30].

Levels of involvement are often used to assess the depth of participation in IT design. Most of the included studies involved nurses as users and testers of technologies – usually as research participants [26, 202]. Consistent with Bos et al. [91] study, nurses were found to engage in testing and providing feedback on health technologies that had already been developed. While providing feedback on technologies is important, most studies mainly sought input from nurses rather than engaging them as co-designers [43, 227, 228]. This consultative role may not only suggest that nurses are not regarded as partners in the health IT design life cycle as recommended by the ISO 9241-210 standards [144, 148] but also a disregard of its significance. Addressing these challenges is key to improving design outcomes. As health IT systems design grows, so does the importance of designing with nurses [30].

Different design techniques were adopted to engage nurses in health IT design, predominantly utilizing participatory user-centered design. However, the concepts of user-centred design and human-centred design were used as equivalent terms and mostly referred to the application of the ISO 9241-210 requirements [144, 148]. User-centred design is considered a design strategy that centres on the needs of the user in health IT design [25] whereas human-centred design involves not only the user’s needs but also the broader context of empathizing with users to understand their needs and challenges [27]. Moreover, a variety of design methods were utilized to engage nurses. Consistent with our findings, Richardson et al. [70] observed that research focusing on usability testing of health IT systems often apply concurrent think-aloud technique and interviews for users to verbalize their thoughts and feelings as they interact with the software. The use of these design approaches and methods are meant to reinforce the need for and importance of meaningfully engaging nurses in the design process [26]. However, it is unclear which design methods or approaches were more effective in engaging nurses in the design process. Thus, further research is needed to examine the effectiveness of design-based methods in engaging nurses in health IT design according to evidence-based standards [30].

Although we did not specifically assess the demographic characteristics of nurses who were included in the design studies, we reported on the number of studies that assessed their personal characteristics such as age and years of experience. Future reviews should not only report on studies that assess nurses’ characteristics but also explore the specific demographics of nurses who are most often involved in the health IT design process. Understanding the demographics of nurses who are often involved in health IT design could advance equity and inclusive design [225]. Consistent with an earlier study by Cui et al. [128], nurses’ demographic characteristics, including age, gender and years of experience can influence their levels of engagement and readiness to accept health IT tools after deployment. Our findings therefore suggest that software developers and hospital managers could motivate nurses with diverse backgrounds and expertise to actively participate in the design of health IT systems for patient care.

### Policy, practice, educational, and research implications

The findings of this review highlight the need for hospital managers and policy-makers to prioritize nurse involvement in design by allocating time for their participation. Ongoing training and educational opportunities need to be offered to nurses to improve upon their health IT design skills. Additionally, hospital boards could establish team-based design groups to facilitate co-creation of health IT systems with the nurse in the forefront.

The late-stage involvement of nurses we identified herein stresses that nurses’ inclusion remains tokenistic, despite the recommendations by the ISO 9241-210. Therefore, in practice, direct care nurses could be engaged early and at the upstream design stages in creating and customizing health IT systems that align with patient care. Furthermore, our findings suggest the need to identify and incorporate necessary competencies, requirements, and roles of nurses in health IT design in nursing schools’ curricula. For instance, Van Houwelingen et al. [30] recommend for education, training, and upskilling opportunities to equip nurses in direct care with the skills to innovate and design health IT systems for use in clinical practice.

Lastly, our study underscores the need for further research on design methods and approaches that are most effective in engaging nurses according to evidence-based standards. Future research should also focus on exploring how nurses prefer to be involved in health IT design, factors that influence their engagement, and the specific contributions and roles they can play in designing, developing, or deploying health IT systems. Understanding these approaches and roles is critical for effective participation of nurses in health IT design that align with the realities of nursing practice.

## Limitations

This review study has limitations that need acknowledgment. Conference abstracts, case reports, commentaries and gray literature that might have contained relevant information to this review study were excluded due to lack of rigorous peer review. However, this exclusion criteria were important in ensuring quality of our findings. No appraisal tools were used to appraise the quality of the included studies and therefore – however this is not required by JBI. Additionally, no validated tools were used to assess the outcome of the design process but solely based on nurses’ feedback reported in the studies. Our search only included studies published in English - this restricted eligibility might have excluded relevant studies published in other languages.

## Conclusions

Our review shows that nurses were frequently included at the later stages of health IT design process with very little opportunity to make contributions to key components of the design.

Additionally, although other methods such as interviews, surveys, card sorting, storyboarding and observations were also used as design methods, it was unclear which methods were most effective in engaging nurses in the design process. Also, no standardized or validated nurse engagement frameworks were found to exist in this review. Nevertheless, the study identified the significant impacts of nurses’ inclusion in design, including increased acceptability, usability and satisfaction with the health IT systems. By indicating the gaps in current research, this review sets a foundation for advancing nurse-centered design in health IT systems research by promoting future nursing engagement in health IT design processes.

## Data Availability

All data are in the manuscript and/or supporting information files.

## Acknowledgements

The authors acknowledge Katherine Miller and Ellis Ursula, Health Science librarians at the University of British Columbia for their support. We also gratefully thank Joanne Callow and Dr. Leanne Currie, an Associated Professor at the School of Nursing, University of British Columbia for their contributions to this project. Dr. Kristen Haase is supported by a Health Services BC Scholar Award.

## Author contributions

**Conceptualization**: Francis Kobekyaa, Kristen Haase

**Formal analysis**: Francis Kobekyaa

**Investigation**: Francis Kobekyaa, Dominique Boswell, Reilly Sinclair

**Methodology**: Francis Kobekyaa

**Software:** Francis Kobekyaa

**Validation:** Kristen Haase

**Visualization:** Francis Kobekyaa

**Supervision**: Kristen Haase, Havaei Farinaz, and Tracie Risling

**Writing – original draft**: Francis Kobekyaa

**Writing – review & editing**: Francis Kobekyaa, Dominique Boswell, Reilly Sinclair, Farinaz Havaei, Tracie Risling, Kristen Haase.

## Supporting information

**S1 Table.** Protocol for the nurse involvement in health information technology design for digital nursing practice.

(DOC)

**S2 Table.** Preferred Reporting Items for Systematic Reviews and Meta-analysis Extension for Scoping Reviews (PRSIMA-ScR) Checklist.

(DOC)

**S3 Table.** Database search strategy

(DOC).

**S4 Table.** Characteristics of each included study.

(DOC).

