## Supplementary material for "Nurse involvement in health information technology design for digital nursing practice: a scoping review": S3 Table

**S3 Table. Database Search Strategy**

| **Database** | **Search** | **Results** |
| --- | --- | --- |
| Ovid MEDLINE(R) <1946 to April 9, 2025> | 1. exp nurse/ | 98675 |
|  | 2. exp Nursing Care/ | 141531 |
|  | 3. exp nursing staff/ | 69874 |
|  | 4. exp nursing/ | 263646 |
|  | 5. nurs*.tw,kf. | 534447 |
|  | 6. 1 or 2 or 3 or 4 or 5 | 718788 |
|  | 7. Software Design/ | 6139 |
|  | 8. user-centered design/ | 195 |
|  | 9. exp user-computer interface/ | 39470 |
|  | 10. exp Universal Design/ | 293 |
|  | 11. (agile software development method* or co-design* or computer graphic* or design thinking process or double diamond model* or end-user engag* or end-user involv* or end-user participat* or graphical user interfac* or human-centered design* or human-centred design* or human-centered comput* or human-centred comput* or participatory design* or software design* or system design* or usability or user-centered comput* or user-centred comput* or user-centered design* or user-centred design* or user-computer interfac* or user co-design* or user involv* or universal design*).tw,kf. | 43346 |
|  | 12. 7 or 8 or 9 or 10 or 11 | 83734 |
|  | 13. exp medical informatics/ | 498623 |
|  | 14. nursing informatics/ | 1640 |
|  | 15. information system/ | 19487 |
|  | 16. information technology/ | 813 |
|  | 17. clinical decision support system/ | 9474 |
|  | 18. clinical decision support system* or computerized medical records system* or electronic health record* or electronic medical record* or electronic nursing record* or graphical user display or information system* or information technolog* or intelligent alerting system* or medical informatics or nursing informatics).tw,kf. | 114984 |
|  | 19. 13 or 14 or 15 or 16 or 17 or 18 | 583852 |
|  | 20. 6 and 12 and 19 | 1576 |
| Embase <1974 to April 9, 2025> | 1. exp nurse/ | 216118 |
|  | 2. exp nursing staff/ | 75864 |
|  | 3. exp nursing care/ | 40442 |
|  | 4. exp nursing/ | 380728 |
|  | 5. nurs*.tw,kf. | 616021 |
|  | 6. 1 or 2 or 3 or 4 or 5 | 836850 |
|  | 7. exp universal design/ | 464 |
|  | 8. software design/ | 1277 |
|  | 9. user computer interface/ | 28927 |
|  | 10. user-centered design/ | 441 |
|  | 11. (agile software development method* or co-design* or computer graphic* or design thinking process or double diamond model* or end-user engag* or end-user involv* or end-user participat* or graphical user interfac* or human-centered design* or human-centred design* or human-centered comput* or human-centred comput* or participatory design* or software design* or system design* or usability or user-centered comput* or user-centred comput* or user-centered design* or user-centred design* or user-computer interfac* or user co-design* or user involv* or universal design*).tw,kf. | 53351 |
|  | 12. 7 or 8 or 9 or 10 or 11 | 79149 |
|  | 13. medical informatics/ | 23202 |
|  | 14. nursing informatics/ | 1713 |
|  | 15. information technology/ | 13471 |
|  | 16. information system/ | 41219 |
|  | 17. clinical decision support system/ | 5853 |
|  | 18. (clinical decision support system* or computerized medical records system* or electronic health record* or electronic medical record* or electronic nursing record* or graphical user display or information system* or information technolog* or intelligent alerting system* or medical informatics or nursing informatics).tw,kf. | 175887 |
|  | 19. 13 or 14 or 15 or 16 or 17 or 18 | 224794 |
|  | 20. 6 and 12 and 19 | 5121 |
| CINAHL Ebscohost | S1. (MH "Nurses+") | 229,797 |
|  | S2. (MH "Nursing Care") | 28,148) |
|  | S3. TI ( nurs* ) OR AB ( nurs* ) | 617,021 |
|  | S4. S1 OR S2 OR S3 | 708,116 |
|  | S5. (MH "Systems Design") or (MH "User-Computer Interface+") | 15,525 |
|  | S6. (MH "Software Design") or (MH "Universal Design") | 4,557 |
|  | S7. TI ( "agile software development method*" OR "co-design*" OR "computer graphic*" OR "design thinking process*" OR "double diamond model*" OR "end-user engag*" OR "end-user involv*" OR "end-user participat*" OR "graphical user interfac*" OR "human-centered design*" OR "human-centred design*" OR "human-centered comput*" OR "human-centred comput*" OR "participatory design*" OR "software design*" OR "system design*" OR "usability" OR "user-centered comput*" OR "user-centred comput*" OR "user-centered design*" OR "user-centred design*" OR "user-computer interfac*" OR "user co-design*" OR "user involv*" OR "universal design*") OR AB ( "agile software development method*" OR "co-design*" OR "computer graphic*" OR "design thinking process*" OR "double diamond model*" OR "end-user engag*" OR "end-user involv*" OR "end-user participat*" OR "graphical user interfac*" OR "human-centered design*" OR "human-centred design*" OR "human-centered comput*" OR "human-centred comput*" OR "participatory design*" OR "software design*" OR "system design*" OR "usability" OR "user-centered comput*" OR "user-centred comput*" OR "user-centered design*" OR "user-centred design*" OR "user-computer interfac*" OR "user co-design*" OR "user involv*" OR "universal design*" ) | 15,097 |
|  | S8. S5 OR S6 OR S7 | 32,148 |
|  | S9. (MH "Information System+") or (MH"information technology+") or (MH "Medical Informatics") OR (MH "Nursing Informatics") or (MH "Decision Support Systems, Clinical") | 55,948 |
|  | S10. S9 OR S10 | 83,898 |
|  | S11. S4 AND S8 AND S11 | 3017 |
| Web of Science | **nurs* (Topic) AND "agile software development method*" OR "co-design*" OR "computer graphic*" OR "design thinking process*" OR "double diamond model*" OR "end-user engag*" OR "end-user involv*" OR "end-user participat*" OR "graphical user interfac*" OR "human-centered design*" OR "human-centred design*" OR "human-centered comput*" OR "human-centred comput*" OR "participatory design*" OR "software design*" OR "system design*" OR "usability" OR "user-centered comput*" OR "user-centred comput*" OR "user-centered design*" OR "user-centred design*" OR "user-computer interfac*" OR "user co-design*" OR "user involv*" OR "universal design*" (Topic) AND "clinical decision support system*" OR "computerized medical records system*" OR "electronic health record*" OR "electronic medical record*" OR "electronic nursing record*" OR "graphical user display*" OR "information technolog*" OR "information system*" OR "intelligent alerting system*" OR "medical informatics" OR "nursing informatics" (Topic)** | 1288 |
| Scopus | ( TITLE-ABS-KEY ( nurse  OR  nursing  OR  "nursing care"  OR  "nursing staff" )  AND  TITLE-ABS-KEY ( "agile software development method*"  OR  "co-design*"  OR  "computer graphic*"  OR  "design thinking process"  OR  "double diamond model*"  OR  "end-user engag*"  OR  "end-user involv*"  OR  "end-user participat*"  OR  "graphical user interfac*"  OR  "human-centered design*"  OR  "human-centred design*"  OR  "human-centered comput*"  OR  "human-centred comput*"  OR  "participatory design*"  OR  "software design*"  OR  "system design*"  OR  usability  OR  "user-centered comput*"  OR  "user-centred comput*"  OR  "user-centered design*"  OR  "user-centred design*"  OR  "user-computer interfac*"  OR  "user co-design*"  OR  "user involv*"  OR  "universal design" )  AND  TITLE-ABS-KEY ( "clinical decision support system*"  OR  "computerized medical records system*"  OR  "electronic health record*"  OR  "electronic medical record*"  OR  "electronic nursing record*"  OR  "graphical user display"  OR  "information system*"  OR  "information technolog*"  OR  "intelligent alerting system*"  OR  "medical informatics"  OR  "nursing informatics" ) )  AND  ( LIMIT-TO ( LANGUAGE ,  "English" ) ) | 1,507   |
| Compendex Engineering Village | (( ((design*) WN KY)) AND ( ((nurs*) WN KY)) AND ( (({medical informatics} OR {medical information systems} OR {nursing informatics}) WN KY))) | 369 |
| IEEE Xplore | ("All Metadata":nurs*) AND ("All Metadata":design*) AND ("All Metadata":medical informatics OR "All Metadata":nursing informatics) | 159 |
